# Genome-wide association studies identify genetic determinants of synucleinopathy biomarkers

**DOI:** 10.64898/2025.12.29.25343142

**Authors:** Emma N. Somerville, Lang Liu, Michael Ta, Hirotaka Iwaki, Konstantin Senkevich, Roy N. Alcalay, Ziv Gan-Or

## Abstract

**Objective:** α-synucleinopathies are clinically and biologically heterogeneous disorders lacking reliable biomarkers to assist with early diagnosis, disease progression, patient stratification, and therapeutic targeting. Genetic variation is known to impact biomarker levels, influencing their utility and interpretation in research and clinical settings. We aimed to identify common genetic modulators of biomarker levels implicated in α-synucleinopathy pathogenesis.

**Methods:** Genome-wide association studies (GWASs) were conducted on 63 CSF, plasma, and urine biomarkers in 581 individuals from the Parkinson’s Progression Markers Initiative (PPMI). Analyses were adjusted for age, sex, disease status, and principal components. PD- and DLB-risk loci associations were separately assessed for each GWAS.

**Results:** We confirm strong associations between urine bis(monoacylglycerol)phosphate (BMP) isoforms and the variants *LRRK2* p.G2019S and *GBA1* p.N370S, while providing support for BMPs use as a *LRRK2*-PD biomarker. CSF Aβ was significantly associated with an APOE ε4 allele, reinforcing its central role in amyloid regulation. Novel associations were detected between CSF ceramide isoforms the *MCF2L2* and *GMNN* loci, and between CSF tau and the *TP63* locus. Multiple PD risk loci, including *MAPT*, *SIPA1L2*, *MCCC1*, and *RAB29*, were associated with lysosomal lipid biomarkers, highlighting pathway-level convergence.

**Interpretation:** The present study reveals established and novel genetic modulators of potential α-synucleinopathy biomarkers, demonstrating that genetic background significantly shapes biomarker levels. These genetic influences should be accounted for when conducting biomarker-based research, clinical trials, or therapeutic development to ensure accurate interpretation and improve their translational relevance.

## Introduction

Accurate and timely diagnosis of α-synucleinopathies, including Parkinson’s disease (PD), dementia with Lewy bodies (DLB), and multiple system atrophy (MSA), remains a major challenge due to overlapping clinical and pathological features between disorders.^1^ A delayed or incorrect diagnosis can limit appropriate treatment and affect patient quality of life. Even after diagnosis, disease management is complicated by considerable heterogeneity in symptom presentation, progression, severity and response to treatment. These limitations underscore the need for additional tools that can complement existing clinical assessments and support both care and therapeutic development. Consistent and reliable biomarkers reflect one such tool by helping give measurable indicators of underlying biological processes, separate disease subtypes, identify individuals at higher risk of disease development, track progression, and enrich clinical trials for therapeutic development through target engagement, effects of disease progression and patient stratification.^2^

Several potential biomarkers are being investigated for clinical and therapeutic applications in α-synucleinopathies. Fittingly, α-synuclein has been the most commonly used and potentially most promising biomarker, specifically in regard to α-synuclein seed amplification assays (SAAs) which have high sensitivity and specificity for diagnosing PD as well as evidence for potentially being able to differentiate PD from DLB and MSA.^3^ Alzheimer’s disease biomarkers (AD) like β-amyloid (Aβ) have also held some promise for differentiating DLB from PD with and without dementia.^4^ Additionally, biomarkers in the glucocerebrosidase (GCase) lysosomal metabolism pathway have been investigated both as therapeutic targets and as indicators for severity of cognitive symptoms and progression.^5, 6^

Biomarker levels and activity can be affected by genetic modifiers, which can complicate their interpretation and utility. For instance, many PD patients that are carriers of biallelic *PRKN* or heterozygous *LRRK2* variants exhibit markedly reduced SAA positivity, illustrating how genetic context can bias biomarker performance and confound both research and clinical trial results.^7^ Understanding these genetic modifiers that affect biomarker profiles can improve their reliability and translational potential, as well as shed light on disease mechanisms. To that end, in the present study we aimed to identify common genetic variants associated with biomarker levels and activities in α-synucleinopathies. Using data from the Parkinson’s Progression Markers Initiative (PPMI) cohort, we analyzed 63 candidate biomarkers to identify potential associations with PD risk and 59 biomarkers with genome-wide association studies (GWASs) to uncover their genetic modulators.

## Methods

### Study Participants

The PPMI cohort included 445 sporadic PD cases, 164 neurologically healthy controls, 51 individuals with parkinsonism and scans without evidence of dopaminergic deficit (SWEDD), and 97 prodromal individuals selected based on symptoms like REM-sleep behaviour disorder (RBD) and first-degree family history of PD. Full cohort inclusion and exclusion criteria can be found on the PPMI website (https://www.ppmi-info.org/study-design/study-cohorts). Some individuals in this cohort are genetically enriched, having been recruited for the presence of known genetic PD-risk variants in *LRRK2*, *GBA1*, and *SNCA*. The PPMI cohort has been described in detail previously.^8, 9^ All participants in this study were of European ancestry, confirmed with principal component analysis (PCA). Informed consent forms were administered and signed by all participants before entering the study, and the study protocol was approved by the relevant institutional review boards.

### Biomarker Collection

Bis(monoacylglycerol)phosphate (BMP) isoforms were extracted from urine aliquots through liquid-liquid extraction and measured using ultra-performance liquid chromatography with tandem mass spectrometry (UPLC-MS/MS), the methodology has been described previously.^10, 11^ The protocol for extraction of lipid metabolites, including ceramides (Cer), lactosylceramides (GL2), and sphingomyelins (SM), has been previously described.^12^ Briefly, the sphingolipids were extracted from plasma by protein precipitation in extraction solution containing internal standards cocktails (<u>PPMI</u>, Project 135). The same sphingolipids were extracted from cerebrospinal fluid (CSF) by liquid-liquid extraction, with the exception of SM which was extracted with protein precipitation. Protein precipitation samples were exposed to an extraction solution containing internal standards (IS), while liquid-liquid extraction samples were suspended with methanol, chloroform, water and ISs. After vortex and centrifugation, quantitative analyses of all sphingolipids were performed using liquid chromatography with tandem mass spectrometry (LC-MS/MS). CSF Aβ, total tau (t-tau), and phosphorylated tau (p-tau) levels were measured using Elecsys electrochemiluminescence (ECL) immunoassays on a cobas e 601 analyzer (Roche Diagnostics) (<u>PPMI</u>, Project 125). Enzyme methodology for additional biomarkers can be found to approved researchers through the USC Laboratory of Neuro Imagine (LONI) Image & Data Archive (IDA) (https://ida.loni.usc.edu/login.jsp). Biomarker mean calculations and linear regressions with PD disease status were performed in R v4.2.1.^13^ Biomarkers with skewed distributions, including CSF tau isoforms, CSF Aβ, CSF α-synuclein (aSyn), urine BMP isoforms, and serum neurofilament light chain (NfL), were log-transformed prior to analyses.

### Statistical Analyses

The PPMI cohort consisted of two genotyping datasets based on the array used: the NeuroX SNP array for the original recruitment cohort consisting of Parkinson’s disease cases, healthy controls, and SWEDD individuals, and the OmniExpress Exome+ v1.3NeuroX array for the more recently recruited cohort consisting largely of prodromal and genetically recruited PD cases and controls. Each dataset was subjected to quality control, imputation, and regression separately, followed by meta-analysis for biomarkers that were measured in both datasets. Quality control for individual-and variant-level data was completed as previously described (https://github.com/neurogenetics/GWAS-pipeline).^14^ Imputation was performed on cleaned data with the TOPMed Imputation Server using the TOPMED reference panel r3 and default settings.^15^ GWASs were performed with linear regression in plink v1.9 for all biomarkers with data in at least 400 individuals, using soft-called variants (R^2^ > 0.3) and a minor allele frequency (MAF) threshold of 0.05.^16^ Adjustments for age, sex, disease status, and the top 5 principal components (PCs) were included. Fixed-effect meta-analyses were performed for relevant biomarkers using METAL in plink v1.9.^17^ Conditional and joint analysis was performed with GCTA-COJO to identify independent associations when adjusting for the lead variant in significant loci.^18^ Linkage disequilibrium (LD) statistics were obtained through LDlink (https://ldlink.nih.gov/?tab=home).^19^ Logistic regressions of biomarker levels with PD disease status and area under the curve (AUC) analyses were performed in R v4.2.1 using age, sex, and the top 5 PCs as covariates. Individuals from the SWEDD and prodromal sub-cohorts were excluded from this analysis.

## Results

Following quality control, 63 biomarkers with 581 individuals and 7,099,634 variants from the NeuroX array and 200 individuals and 15,364,231 variants from the OmniExpress array available for analyses. Biomarkers showing evidence of bias in their GWAS results based on inflated lambdas and atypical Manhattan plots, including CSF α-synuclein, CSF hemoglobin, serum neurofilament light chain, and nuclear DNA beta-2-microglobulin copy number, were excluded from GWAS analyses but retained for logistic regressions evaluating their associations with PD risk, resulting in GWAS analyses of 59 biomarkers overall. QQ plots for markers with notable results are presented here, and lambdas were deemed acceptable (Supplementary Fig 1).

### Biomarker associations with PD risk

Means and medians for each biomarker were calculated and association testing against PD risk was performed after removing SWEDD and prodromal individuals (Supplementary Table 1). In total, 25 of our analyzed biomarkers were nominally associated with PD, with 10 remaining after applying Bonferroni correction (p-value threshold = 6.8e-04). The strongest association was between a positive SAA result and PD disease status. Lower levels of typical AD markers like CSF pTau, tTau, and Ab were associated with PD, as well as a higher Ab/pTau ratio. Higher levels of GL2, SM, and Cer, all part of the lysosomal degradation pathway, were also associated with PD risk. Interestingly, increased total urine di-22:6 and 2,2’-di-22:6 BMP isoforms were both associated with PD, but the total di-18:1 isoform was not. Predictive power was assessed with AUC analyses for biomarkers significantly associated with PD, with SAA being the only marker to be strongly predictive of disease status (Fig 1).

**Fig 1.**
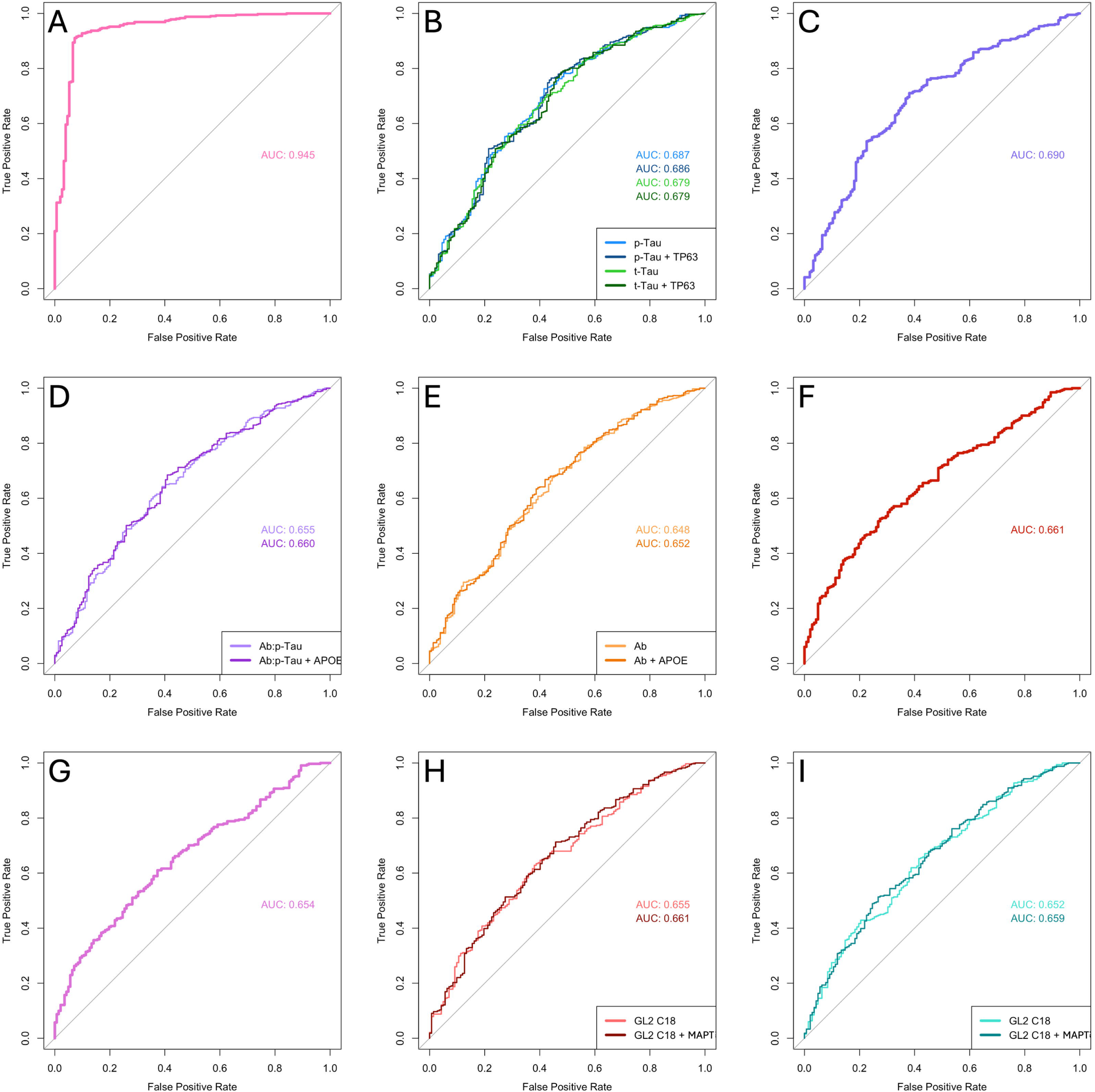
Area under the curve (AUC) analysis for logistic regressions of biomarkers with PD status in the PPMI cohort for A) SAA, B) CSF p-tau and t-tau with and without adjustments for the lead GWAS SNP in *TP63*, C) CSF α-synuclein, D) CSF Aβ:p-tau ratio with and without adjustment for the lead GWAS SNP in *APOE*, E) CSF Aβ with and without adjustment for the lead GWAS SNP in *APOE*, F) total CSF GL2, G) total plasma GL2, H) plasma C18 GL2 with and without adjustment for the lead GWAS SNP in *MAPT*, and I) plasma C16 GL2 with and without adjustment for the lead GWAS SNP in *MAPT*.

### Genetic modulators of biomarker levels

We identified notable associations for multiple biomarkers. First, we observed that the 2,2’-di-22:6, total di-18:1, and total di-22:6 BMP isoforms were positively associated with *LRRK2* p.G2019S and negatively associated with signals in *GBA1* likely driven by p.N370S (Fig 2, Table 1). No secondary independent signals were identified in either locus after conditional analysis. The *GBA1* association remained significant after adjustment for *LRRK2* p.G2019S in the 2,2’-di-22:6 and total di-22:6 BMP analyses, but not in the total di-18:1 BMP analysis (Supplementary Fig 2). These findings were supported by replication in an external cohort from Columbia University, where creatinine-normalized di-18:1-BMP levels were significantly lower in *GBA1* carriers (n=18) compared to non-carriers (n=57) (*GBA1*+, median = 1.15, range = 0.0-3.86; *GBA1*-, median = 1.39, range = 0.0-12.7; p= 0.0259).^20^ The same trend was observed for di-22:6-BMP, although it did not reach statistical significance (*GBA1*+, median = 4.29, range = 1.15-22.17; *GBA1*-, median = 5.62, range = 0.0-25.8; p= 0.0619). Notably, we did not replicate previous findings of a significant difference^20^ in BMP levels between *LRRK2* carriers with PD and non-manifesting controls, and did not observe a significant difference within *GBA1* carriers or *LRRK2* and *GBA1* non-carriers when comparing PD cases to controls (Supplementary Fig 3). Our GWASs of CSF ceramides identified the *MCF2L2* locus as associated with the C22, C23, and C24 isoforms (Fig 3, Table 2). The *GMNN* locus was also associated with C24 ceramide levels and showed a near-significant association with the C22 and C23 isoforms. Notably, *MCF2L2* is part of a PD-risk locus comprising *MCF2L2*, *MCCC1*, and *LAMP3*. However, the lead variant in our analysis was not in strong LD with the lead variant in this locus from the Nalls et al. (2019) PD GWAS^21^ (R2 = 0.0039, D’ = 0.62). In our GWAS of CSF Aβ, we identified a strong negative association with the *APOE* locus, driven by one of the e4 haplotype alleles (rs429358, beta = −0.11, se = 0.013, p = 2.62e-15, Fig 4A). Conditional analysis revealed no secondary independent associations. Analysis of CSF t-tau and p-tau levels revealed a negative association between t-tau levels and an intergenic locus near *TP63* (rs1424826, beta = −0.087, se = 0.015, p = 9.17e-09, Fig 4B). This same locus demonstrated a similar trend towards genome-wide significance for CSF p-tau, with a consistent direction and magnitude of effect as what was observed for t-tau (beta = −0.085. se = 0.016, p = 7.23-08). Lastly, our GWASs of Aβ/t-tau and Aβ/p-tau ratios identified associations with *APOE* consistent with those observed for CSF Aβ alone (Ab/pTau, rs429358, b = −0.12, se = 0.02, p = 1.94e-10; Ab/tTau, rs429358, b = −0.054, se = 0.0053, p = 2.5e-24, Fig 4C and D). No association was observed between these ratios and the *TP63* locus observed in the tau analyses. Conditional analyses revealed no secondary independent signals within the *APOE* region.

**Fig 2.**
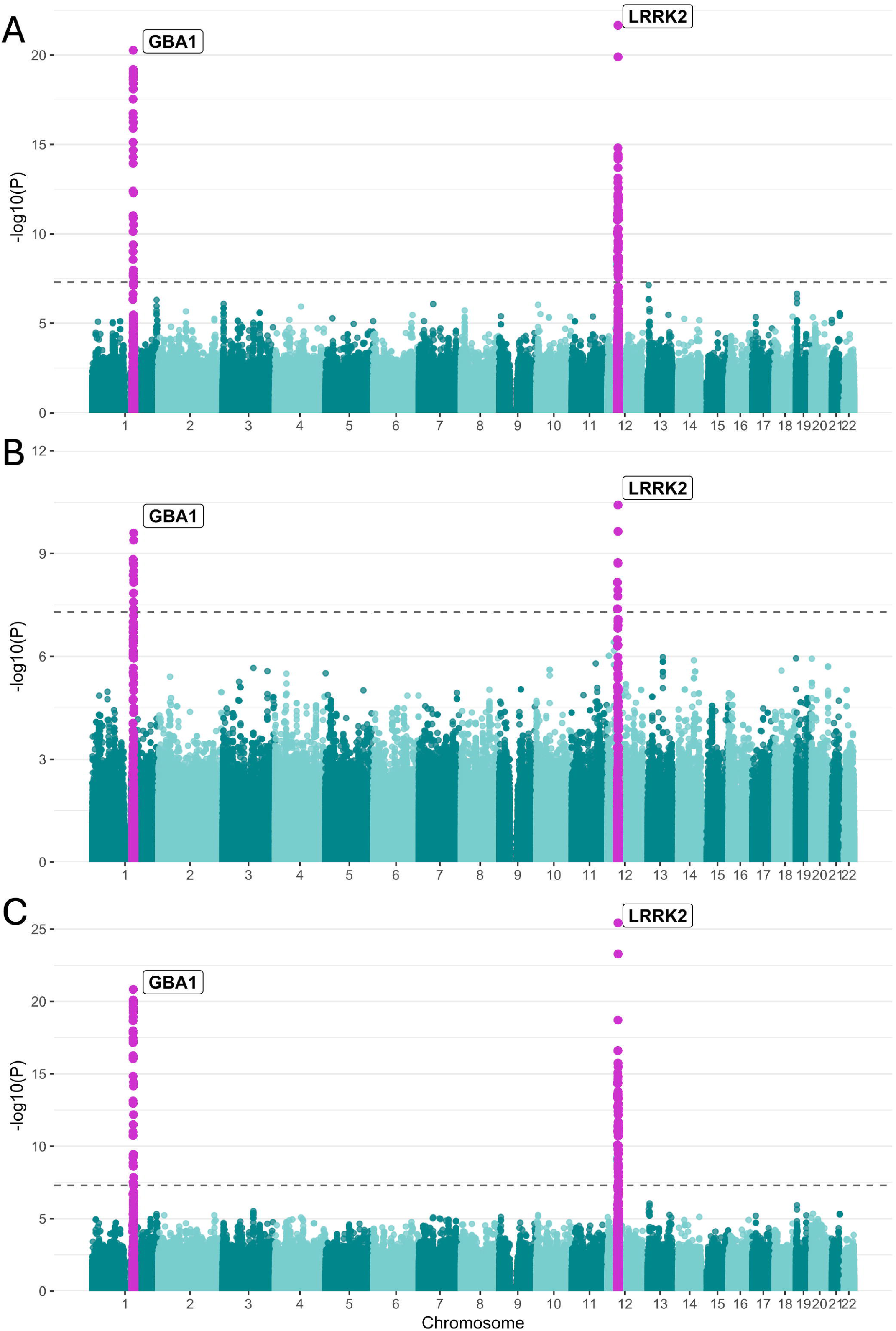
Manhattan plot of log adjusted p-values at each genomic position for GWASs of urine BMP A) 2,2’-di-22:6, B) total di-18:1, and C) total di-22:6 isoforms adjusted for age, sex, disease status, and the top 5 principal components.

**Fig 3.**
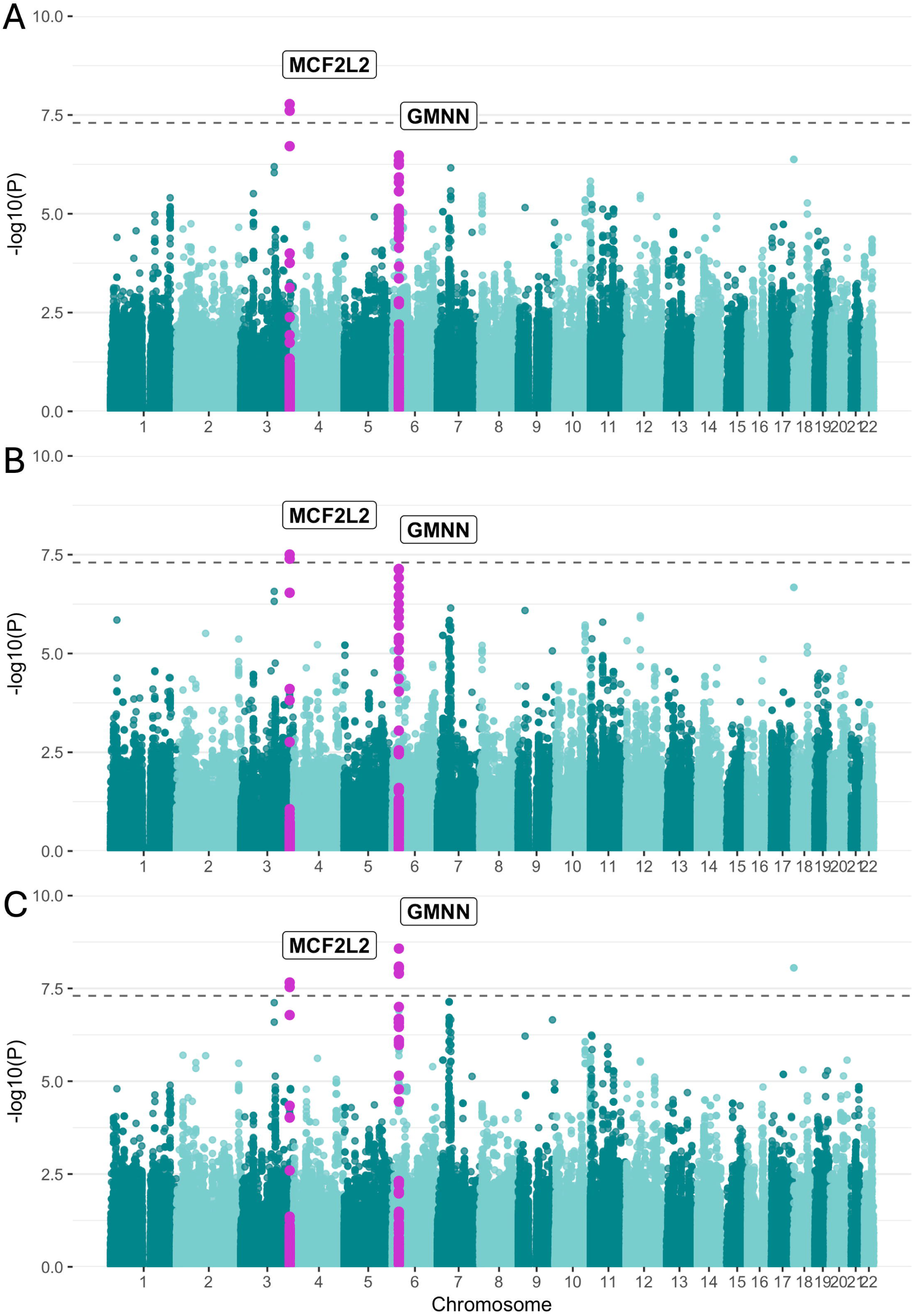
Manhattan plot of log adjusted p-values at each genomic position for GWASs of CSF ceramide A) C22, B) C23, and C) C24 isoforms adjusted for age, sex, disease status, and the top 5 principal components.

**Fig 4.**
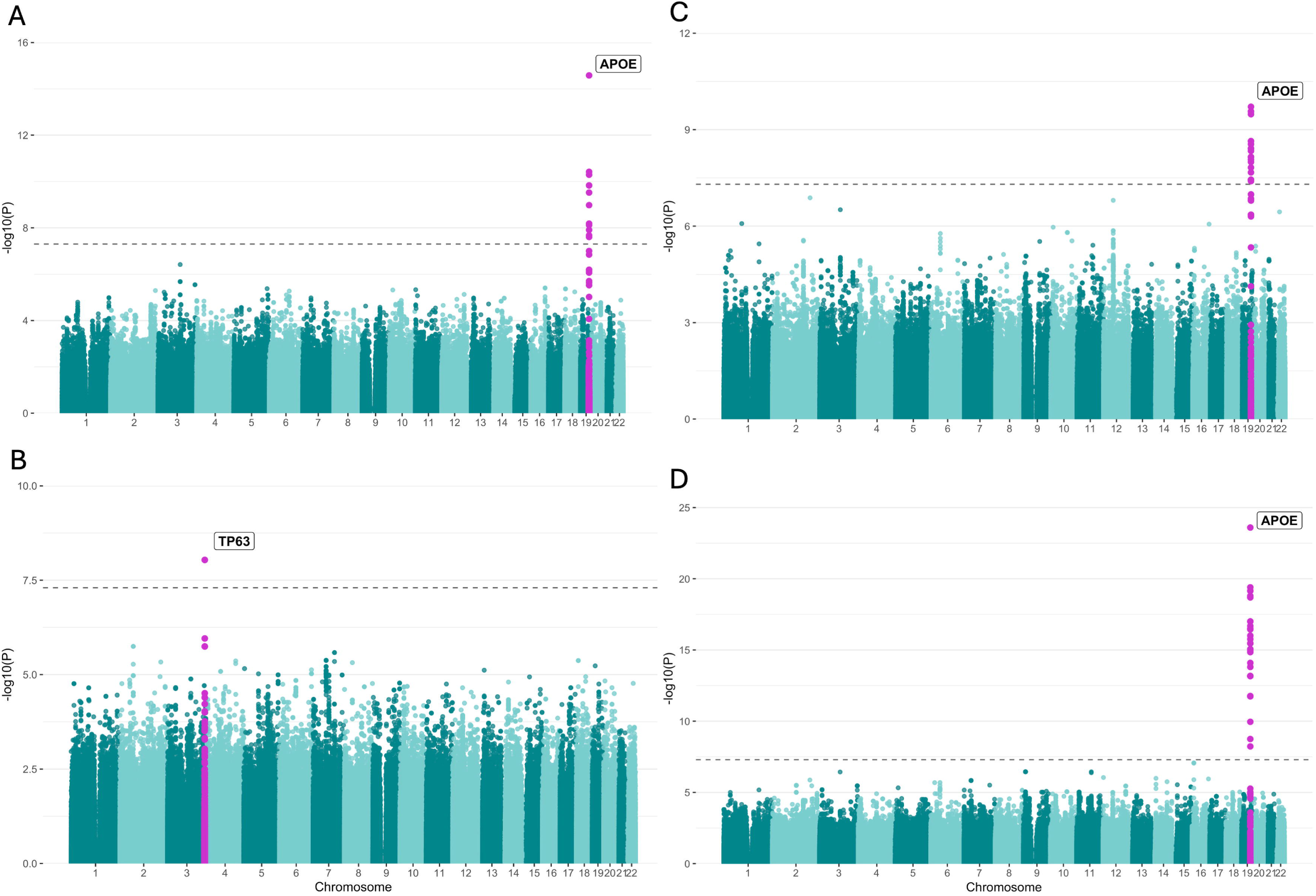
Manhattan plot of log adjusted p-values at each genomic position for GWASs of A) CSF Aβ, B) CSF t-tau, C) Aβ/p-tau, and D) Aβ/t-tau adjusted for age, sex, disease status, and the top 5 principal components.

**Table 1.** Lead *LRRK2* and *GBA1* variants for urine bis(monoacylglycerol)phosphate isoforms from genome-wide association studies.

| **Isoform** | **Gene** | **Lead SNP** | **Beta** | **SE** | **P** |
| --- | --- | --- | --- | --- | --- |
| 2,2'-di-22:6 | *LRRK2* | rs189637182 | 0.6 | 0.061 | 2.17e-22 |
|  | *GBAI* | rs144115539 | -0.46 | 0.049 | 5.41e-21 |
| total di-18:1 | *LRRK2* | rs189637182 | 0.47 | 0.071 | 3.85e-11 |
|  | *GBAI* | rs150850629 | -0.42 | 0.067 | 2.52e-10 |
| total di-22:6 | *LRRK2* | rs189637182 | 0.55 | 0.052 | 3.76e-26 |
|  | *GBAI* | rs61732802 | -0.6 | 0.048 | 1.47e-21 |

SNP, single nucleotide polymorphism; SE, standard error.

**Table 2.** Lead *MCF2L2* and *GMNN* variants for CSF ceramide isoforms from genome-wide association studies.

| **Isoform** | **Locus** | **Lead SNP** | **Beta** | **SE** | **P** |
| --- | --- | --- | --- | --- | --- |
| C22 | *MCF2L2* | rs9819356 | 0.18 | 0.031 | 1.67e-08 |
|  | *GMNN* | rs2817735 | 0.16 | 0.03 | 3.33e-07 |
| C23 | *MCF2L2* | rs9819356 | 0.18 | 0.032 | 3.13e-08 |
|  | *GMNN* | rs75469557 | 0.16 | 0.03 | 7.2e-08 |
| C24 | *MCF2L2* | rs9819356 | 0.49 | 0.087 | 2.18e-08 |
|  | *GMNN* | rs2817735 | 0.51 | 0.083 | 2.68e-09 |

SNP, single nucleotide polymorphism; SE, standard error; CSF, cerebrospinal fluid.

### PD risk variants are associated with lysosomal lipid levels

We investigated whether genetic loci previously associated with PD or DLB influence biomarker levels in our cohort. Using risk variants from the Nalls et al. (2019) PD GWAS^21^ and the Chia et al. (2021) DLB GWAS^22^, we identified 18 associations between PD loci and GL2, Cer, or SM isoform levels, as well as 3 associations between DLB loci and Ab or tau biomarkers after Bonferroni correction (Table 3; PD threshold p < 4.68e-4; DLB threshold p < 1e-02). Among PD loci, the *MAPT* locus was associated with increased levels of plasma GL2 C16, C18, and C23 isoforms. Plasma GL2 C23 levels were also positively associated with a variant in *SIPA1L2*. The *MCCC1;LAMP3* locus showed a positive association with plasma SM C24 isoform levels, while the *RAB29* risk locus was associated with increased levels of plasma Cer C16 and C22 isoforms. We additionally observed associations between variants near *LRRK2* and both 2,2’-di-22:6 and total di-22:6 BMP. However, these signals were not independent in conditional analyses and did not remain significant after adjusting for *LRRK2* p.G2019S carrier status (2,2’-di-22:6, rs117073808, p = 0.65; 2,2’-di-22:6, rs138017112, p = 0.24; total di-22:6, rs117073808, p= 0.21; total di-22:6, rs138017112, p = 0.84), indicating that these associations were likely driven by the p.G2019S variant. In the DLB analysis, the DLB-associated *APOE* variant demonstrate significant associations with decreased CSF Aβ levels, as well as decreased Aβ/p-tau and Aβ/t-tau ratios. This variant is in LD with the lead variant in our analyses (D’ = 1.0, R^2^ = 0.77), meaning these are driven by the same signal.

**Table 3.**
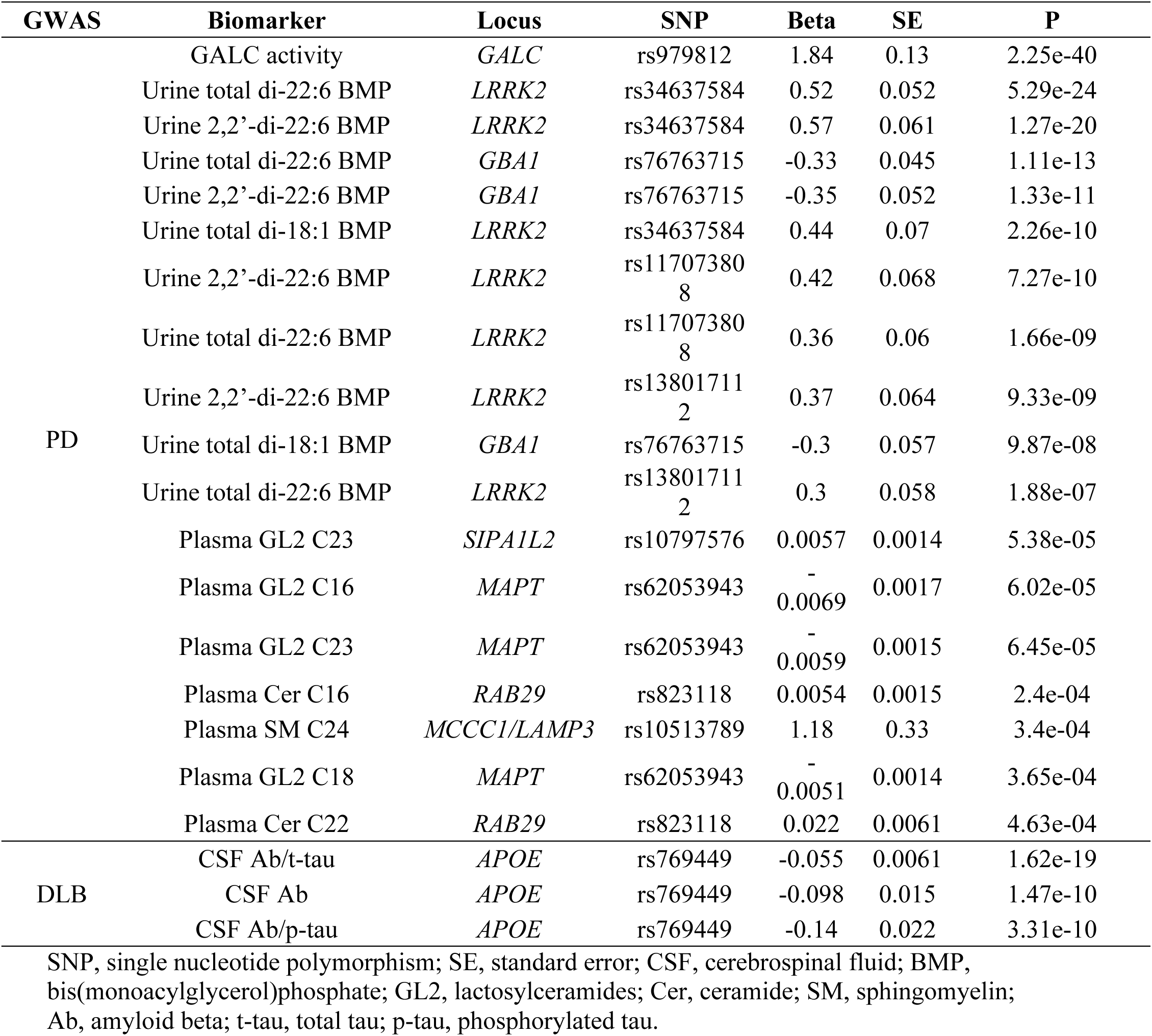
PD and DLB genome-wide association study loci associated with Parkinson’s Progression Markers Initiative biomarkers.

## Discussion

In the present study, we conducted logistic regressions with disease status for 63 biomarkers and GWASs for 59 biomarkers implicated in the pathogenesis of α-synucleinopathies to identify common genetic variants that modulate their levels and activities. By integrating genetic architecture with biomarker data, we provide genetic context for biomarker associations with PD, DLB, and related disorders, and identify genetic modulators that account for significant biomarker variability, enabling improved adjustment and stratification in future research involving these biomarkers.

We found *LRRK2* p.G2019S and *GBA1* p.N370S to be the strongest genetic modifiers of urine BMP isoforms. BMP has been previously suggested as a candidate biomarker for *LRRK2*-PD, supported by evidence of elevated BMP levels in *LRRK2* p.G2019S carriers, correlations with PD status in these individuals, and associations with cognitive decline in a study utilizing Columbia University and LRRK2 Cohort Consortium cohorts (LCC).^20^ In our study, we replicated the association with the *LRRK2* locus in the PPMI cohort. We observed a negative association with *GBA1* p.N370S in two independent cohorts, even when *LRRK2* p.G2019S is adjusted for, in contrast with a previous report suggesting a positive association.^11^ Our results do not support BMP isoforms being associated with PD disease status after multiple testing correction, adding complexity to BMP’s use as a biomarker. Moreover, when comparing cases and controls within *LRRK2* p.G2019S or *GBA1* p.N370S carriers, none of the groups demonstrated significant differences in BMP levels. Together these results indicate that BMP levels are robustly driven by *LRRK2* and *GBA1* genotypes, but do not show consistent disease-state differences, confirming previous suggestions that BMP may serve better as a LRRK2 pathway indicator rather than as a diagnostic biomarker.^20, 23, 24^ Next, we replicated a well-established association at the *APOE* locus for CSF Aβ, specifically identifying a variant defining the ε4 haplotype as the strongest genetic driver of Aβ levels. In addition to its central role in lipid transport, *APOE* has also been shown to regulate Aβ aggregation and clearance.^25, 26^ Accordingly, it is not unexpected that this locus is the strongest regulator of CSF Aβ, with the observed negative direction of effect likely reflecting decreased free Aβ in the CSF due to increased aggregation or amyloid plaque deposition, consistent with previous findings.^27^ Our analyses of CSF ceramide revealed significant associations between the C22, C23, and C24 isoforms and two loci: *MCF2L2* and *GMNN*. The association at the *MCF2L2* locus is particularly notable, as this *MCCC1;LAMP3;MCF2L2* region has previously been linked to reduced PD risk across European, Asian, and Han Chinese populations,^21, 28, 29^ although the lead SNP for this locus is not in LD with the PD-associated variants.^21^ Ceramide dysregulation has been consistently observed in PD and other α-synucleinopathies, and could serve as a marker of lysosomal dysfunction^30^; therefore, the associations reported here may support a mechanistic connection between the *MCCC1;LAMP3;MCF2L2* locus and lysosomal impairment. We additionally detected a novel association with the *GMNN* locus, a gene encoding geminin, which plays a critical role in cell cycle regulation by inhibiting DNA replication.^31^ *GMNN* has not been connected to α-synucleinopathies in previous research and the cause of its association with CSF ceramides is not obvious based on the current literature, therefore further research will be required to validate the association and understand the mechanism behind this link. We identified an association between CSF t-tau levels and the *TP63* locus, while also observing a suggestive association with CSF p-tau. To our knowledge, this region has not been previously implicated in tau biology or tauopathy-related pathology, meaning *TP63* may represent a novel genetic modulator of CSF tau levels. A prior meta-analysis of CSF tau in PPMI and additional cohorts identified an association the nearby *GEMC1;OSTN* locus^8^; however, the lead variant in that study is not in LD with the lead variant presented here, indicating our signal is likely independent and may reflect a distinct biological mechanism. *TP63* encodes tumor protein p63, a member of the p53 transcription factor family, and plays an important role in proliferation and differentiation, regulation of chromatin-modulating factors, DNA damage response, and cell death and survival.^32^ Although the precise mechanism is not yet established, p63 isoforms have been shown to transcriptionally induce tau phosphorylation HEK293a cells at AD-relevant epitopes,^33^ suggesting that variation at *TP63* could contribute to tau dysregulation through p53-family mediated neurodegenerative pathways. Lastly, our GWASs of the Aβ/t-tau and Aβ/p-tau ratios revealed associations with the same *APOE* variant observed for CSF Aβ alone. These findings indicate that the genetic regulation of the Aβ/tau ratios is predominantly influenced by the *APOE* ε4 allele. In contrast, no association was detected at the *TP63* locus identified in the tau-only analyses, suggesting that the *TP63*-mediated effects on tau are not sufficiently large enough to influence Aβ/tau ratios at the current power level and may not be impactful enough to require consideration in future Aβ/tau ratio applications.

Our analyses of PD loci revealed associations between risk variants and multiple lysosomal biomarkers, including GL2, Cer, and SM. The top association in the locus has been thoroughly examined in a previous study.^14^ The *MAPT* locus, encoding tau, was associated with increased plasma levels of CSF GL2 C16, C18 and C23 isoforms. This variant is representative of the H2 haplotype^34^ which is linked to decreased PD risk.^21^ *MAPT* and tau have both been implicated in lysosomal dysfunction in previous studies,^35, 36^ but more specifically, it has also been reported to be linked to increased GL2 levels in a tau transgenic mouse model,^37^ supporting the genetic association we observe in the present study. We also observed an association with *SIPA1L2* and increased plasma GL2 C23 levels. *SIPA1L2* is a member of the signal-induced proliferation-associated 1 like family and is involved in Rap GTPase activation, and has been shown to play a role in amphisome trafficking.^38, 39^ Next, the *MCCC1/LAMP3* locus was associated with decreased plasma SM C24. *MCCC1* encodes the α-subunit of 3-methylcrotonyl-CoA carboxylase, a mitochondrial enzyme involved in leucine catabolism,^40^ while *LAMP3* encodes lysosomal-associated membrane protein 3, which has been implicated in the unfolded protein response and lysosomal metabolism.^41^ Although it is unclear which gene underlies the association of this locus with SM levels, the signal suggests potential crosstalk between mitochondrial metabolism pathways and sphingomyelin regulation, or alternatively, points to a broader role for lysosomal dysfunction in shaping sphingolipid metabolism. Lastly, we found an association between *RAB29* and decreased plasma Cer C16 and C22 isoforms. The protein Rab29 is a substrate of LRRK2 kinase^42^, and variants in *RAB29* and *LRRK2* have been shown to have an epistatic effect on PD risk in previous studies.^43, 44^ The association of this locus with ceramides suggest that this pathway may be mechanistically linked to ceramide metabolism downstream of *LRRK2*, although the exact cause of this is unclear from the current literature. While these associations suggest potential biological mechanisms linking PD risk loci to various aspects of lysosomal metabolism, functional studies are currently lacking, and further research is needed to elucidate their underlying mechanisms.

Our study has several limitations. First, the availability of cohorts with comprehensive biomarker datasets is limited, restricting our ability to replicate the associations identified in this manuscript. We are also limited by the number of individuals within our cohort who have both genetic and biomarker data, reducing our statistical power to detect associations with rare variants or those with smaller effect sizes. These limitations underscore the need for larger, well-phenotyped cohorts that systematically collect both genetic data and biomarker measurements, and the analyses presented here should be revisited to validate and refine findings when such datasets become available. Additionally, many of our biomarkers with significant associations reflect CSF levels, which may not be fully representative of processes occurring in brain tissue or peripheral systems. Our study was restricted to individuals of European ancestry due to reduce population stratification bias, which limits generalizability and underscores the importance of conducting similar studies in diverse populations when data becomes available. Finally, the cross-sectional GWAS design of our analyses precludes conclusions about causality or temporal changes in biomarker levels over the disease course.

In summary, this study analyzed genetic modifiers of numerous biomarkers implicated in α-synucleinopathies, revealing multiple loci that influence lysosomal lipids, amyloid, tau, and BMP levels. Our findings highlight that common variants, particularly in *LRRK2*, *GBA1*, and *APOE*, exert strong effects on biomarker levels and may account for variability in disease manifestation and biomarker stratification. Notably, we also identified several novel biomarker associated loci, such as *TP63* and *GMNN*, which may represent previously unrecognized biological pathways relevant to neurodegeneration. Together, these results underscore the importance of incorporating genetic context into biomarker interpretation. As biomarker measurements becoming increasingly used in research, therapeutic development, and clinical care, it is essential to account for genetic influences that can meaningfully shift their baseline levels and obscure true disease or treatment related signals. Identifying and adjusting for these modulators will be critical for the development of robust, reliable, and actionable biomarkers, as well as for advancing genetically informed precision medicine approaches for PD, DLB, and related disorders.

## Acknowledgments

We gratefully thank the research participants for contributing to this study. We thank Meron Teferra for her assistance. This study was financially supported through grants from the Michael J. Fox Foundation (MJFF, No. 020700), the Canadian Consortium on Neurodegeneration in Aging (CCNA, No. 049–14), and the Canada First Research Excellence Fund (CFREF, No. 247908), awarded to McGill University for the Healthy Brains for Healthy Lives initiative (HBHL). Additionally, the G-Can (GBA1-Canada) Initiative, an open-science collaborative initiative aimed at addressing GBA1 mutation-based Parkinson’s disease, has made contributions to this research. G-Can is supported by The Hilary & Galen Weston Foundation, Silverstein Foundation, and J. Sebastian van Berkom and Ghislaine Saucier. Data used in the preparation of this article were obtained from the Parkinson’s Progression Markers Initiative (PPMI) database (www.ppmi-info.org/access-data-specimens/download-data), RRID:SCR_006431. For up-to-date information on the study, visit www.ppmi-info.org. PPMI – a public-private partnership – is funded by the Michael J. Fox Foundation for Parkinson’s Research, and funding partners; including 4D Pharma, Abbvie, AcureX, Allergan, Amathus Therapeutics, Aligning Science Across Parkinson’s, AskBio, Avid Radiopharmaceuticals, BIAL, BioArctic, Biogen, Biohaven, BioLegend, BlueRock Therapeutics, Bristol-Myers Squibb, Calico Labs, Capsida Biotherapeutics, Celgene, Cerevel Therapeutics, Coave Therapeutics, DaCapo Brainscience, Denali, Edmond J. Safra Foundation, Eli Lilly, Gain Therapeutics, GE HealthCare, Genentech, GSK, Golub Capital, Handl Therapeutics, Insitro, Jazz Pharmaceuticals, Johnson & Johnson Innovative Medicine, Lundbeck, Merck, Meso Scale Discovery, Mission Therapeutics, Neurocrine Biosciences, Neuron23, Neuropore, Pfizer, Piramal, Prevail Therapeutics, Roche, Sanofi, Servier, Sun Pharma Advanced Research Company, Takeda, Teva, UCB, Vanqua Bio, Verily, Voyager. Z.G.O. is supported by the Fonds de recherche du Québec—Santé (FRQS) Chercheurs-Boursiers award and is a William Dawson Scholar. K.S. is supported by a post-doctoral fellowship from the Canada First Research Excellence Fund (CFREF), awarded to McGill University for the Healthy Brains for Healthy Lives initiative (HBHL). E.N.S. gratefully acknowledges funding support for this research from Parkinson Canada. This work was supported in part by the Intramural Research Program of the National Institute on Aging (NIA), and the Center for Alzheimer’s and Related Dementias (CARD), within the Intramural Research Program of the National Institute on Aging and the National Institute of Neurological Disorders and Stroke (1ZIA-AG000546). The contributions of the NIH authors are considered Works of the United States Government. The findings and conclusions presented in this paper are those of the authors and do not necessarily reflect the views of the NIH or the U.S. Department of Health and Human Services.

## Author Contributions

E.N.S., M.T., H.I., and Z.G.O. contributed to the conception and design of the study. All authors contributed to the acquisition and analysis of data. E.N.S. and Z.G.O. contributed to drafting a significant portion of the manuscript or figures.

## Data Availability

Data used in the preparation of this article were obtained from the Parkinson’s Progression Markers Initiative (PPMI) database (www.ppmi-info.org/access-data-specimens/download-data), RRID:SCR_006431. For up-to-date information on the study, visit www.ppmi-info.org. GWAS summary statistics can be found on the GWAS catalog (https://www.ebi.ac.uk/gwas/). All code used in the present study can be found at https://github.com/gan-orlab/PPMI-Biomarkers-GWASs/tree/main.

## References

1. Tolosa E, Garrido A, Scholz SW, Poewe W. Challenges in the diagnosis of Parkinson’s disease. Lancet Neurol. 2021 May;20(5):385–97.

2. Yamashita KYB, S.; Silverglate, B. D.; Grossberg, G. T. Biomarkers in Parkinson’s disease: A state of the art review. Biomarkers in Neuropsychiatry. 2023;9:100074.

3. Concha-Marambio L, Pritzkow S, Shahnawaz M, Farris CM, Soto C. Seed amplification assay for the detection of pathologic alpha-synuclein aggregates in cerebrospinal fluid. Nat Protoc. 2023 Apr;18(4):1179–96.

4. Hepp DH, Vergoossen DL, Huisman E, et al. Distribution and Load of Amyloid-beta Pathology in Parkinson Disease and Dementia with Lewy Bodies. J Neuropathol Exp Neurol. 2016 Oct;75(10):936–45.

5. Giladi N, Alcalay RN, Cutter G, et al. Safety and efficacy of venglustat in GBA1-associated Parkinson’s disease: an international, multicentre, double-blind, randomised, placebo-controlled, phase 2 trial. Lancet Neurol. 2023 Aug;22(8):661–71.

6. Gonzalez MC, Oftedal L, Lange J, et al. Relationship of cognitive decline with glucocerebrosidase activity and amyloid-beta 42 in DLB and PD. Ann Clin Transl Neurol. 2025 May;12(5):915–24.

7. Hoglinger GU, Adler CH, Berg D, et al. A biological classification of Parkinson’s disease: the SynNeurGe research diagnostic criteria. Lancet Neurol. 2024 Feb;23(2):191–204.

8. Ta M, Blauwendraat C, Antar T, et al. Genome-Wide Meta-Analysis of Cerebrospinal Fluid Biomarkers in Alzheimer’s Disease and Parkinson’s Disease Cohorts. Mov Disord. 2023 Sep;38(9):1697–705.

9. Marek K, Chowdhury S, Siderowf A, et al. The Parkinson’s progression markers initiative (PPMI) - establishing a PD biomarker cohort. Ann Clin Transl Neurol. 2018 Dec;5(12):1460–77.

10. Liu N, Tengstrand EA, Chourb L, Hsieh FY. Di-22:6-bis(monoacylglycerol)phosphate: A clinical biomarker of drug-induced phospholipidosis for drug development and safety assessment. Toxicol Appl Pharmacol. 2014 Sep 15;279(3):467–76.

11. Merchant KM, Simuni T, Fedler J, et al. LRRK2 and GBA1 variant carriers have higher urinary bis(monacylglycerol) phosphate concentrations in PPMI cohorts. NPJ Parkinsons Dis. 2023 Feb 28;9(1):30.

12. Somerville EN, James A, Beetz C, et al. Plasma glucosylceramide levels are regulated by ATP10D and are not involved in Parkinson’s disease pathogenesis. medRxiv. 2024 Sep 16.

13. R Development Core Team. R: A language and environment for statistcal computing. 4.3.1 ed. Vienna, Austria: R Foundation for Statistical Computing; 2023.

14. Senkevich K, Zorca CE, Dworkind A, et al. GALC variants affect galactosylceramidase enzymatic activity and risk of Parkinson’s disease. Brain. 2023 May 2;146(5):1859–72.

15. Taliun D, Harris DN, Kessler MD, et al. Sequencing of 53,831 diverse genomes from the NHLBI TOPMed Program. Nature. 2021 Feb;590(7845):290–9.

16. Purcell S, Neale B, Todd-Brown K, et al. PLINK: a tool set for whole-genome association and population-based linkage analyses. Am J Hum Genet. 2007 Sep;81(3):559–75.

17. Willer CJ, Li Y, Abecasis GR. METAL: fast and efficient meta-analysis of genomewide association scans. Bioinformatics. 2010 Sep 1;26(17):2190–1.

18. Yang J, Ferreira T, Morris AP, et al. Conditional and joint multiple-SNP analysis of GWAS summary statistics identifies additional variants influencing complex traits. Nat Genet. 2012 Mar 18;44(4):369–75, S1-3.

19. Machiela MJ, Chanock SJ. LDlink: a web-based application for exploring population-specific haplotype structure and linking correlated alleles of possible functional variants. Bioinformatics. 2015 Nov 1;31(21):3555–7.

20. Alcalay RN, Hsieh F, Tengstrand E, et al. Higher Urine bis(Monoacylglycerol)Phosphate Levels in LRRK2 G2019S Mutation Carriers: Implications for Therapeutic Development. Mov Disord. 2020 Jan;35(1):134–41.

21. Nalls MA, Blauwendraat C, Vallerga CL, et al. Identification of novel risk loci, causal insights, and heritable risk for Parkinson’s disease: a meta-analysis of genome-wide association studies. Lancet Neurol. 2019 Dec;18(12):1091–102.

22. Chia R, Sabir MS, Bandres-Ciga S, et al. Genome sequencing analysis identifies new loci associated with Lewy body dementia and provides insights into its genetic architecture. Nat Genet. 2021 Mar;53(3):294–303.

23. Gomes S, Garrido A, Tonelli F, et al. Elevated urine BMP phospholipids in LRRK2 and VPS35 mutation carriers with and without Parkinson’s disease. NPJ Parkinsons Dis. 2023 Apr 4;9(1):52.

24. Rideout HJ, Chartier-Harlin MC, Fell MJ, et al. The Current State-of-the Art of LRRK2-Based Biomarker Assay Development in Parkinson’s Disease. Front Neurosci. 2020;14:865.

25. Wisniewski T, Drummond E. APOE-amyloid interaction: Therapeutic targets. Neurobiol Dis. 2020 May;138:104784.

26. Kanekiyo T, Xu H, Bu G. ApoE and Abeta in Alzheimer’s disease: accidental encounters or partners? Neuron. 2014 Feb 19;81(4):740–54.

27. Hashimoto T, Serrano-Pozo A, Hori Y, et al. Apolipoprotein E, especially apolipoprotein E4, increases the oligomerization of amyloid beta peptide. J Neurosci. 2012 Oct 24;32(43):15181–92.

28. Li NN, Tan EK, Chang XL, et al. MCCC1/LAMP3 reduces risk of sporadic Parkinson’s disease in Han Chinese. Acta Neurol Scand. 2013 Aug;128(2):136–9.

29. Sharma M, Ioannidis JP, Aasly JO, et al. Large-scale replication and heterogeneity in Parkinson disease genetic loci. Neurology. 2012 Aug 14;79(7):659–67.

30. Vos M, Klein C, Hicks AA. Role of Ceramides and Sphingolipids in Parkinson’s Disease. J Mol Biol. 2023 Jun 15;435(12):168000.

31. Burrage LC, Charng WL, Eldomery MK, et al. De Novo GMNN Mutations Cause Autosomal-Dominant Primordial Dwarfism Associated with Meier-Gorlin Syndrome. Am J Hum Genet. 2015 Dec 3;97(6):904–13.

32. Fisher ML, Balinth S, Mills AA. p63-related signaling at a glance. J Cell Sci. 2020 Sep 11;133(17).

33. Hooper C, Meimaridou E, Tavassoli M, Melino G, Lovestone S, Killick R. p53 is upregulated in Alzheimer’s disease and induces tau phosphorylation in HEK293a cells. Neurosci Lett. 2007 May 11;418(1):34–7.

34. Senkevich K, Bandres-Ciga S, Cisterna-Garcia A, et al. Genome-wide association study stratified by MAPT haplotypes identifies potential novel loci in Parkinson’s disease. medRxiv. 2023 Apr 15.

35. Somerville EN, Krohn L, Senkevich K, et al. Genome-Wide Association Study of Glucocerebrosidase Activity Modifiers. Mol Neurobiol. 2025 Apr 29.

36. Hung C, Patani R. Elevated 4R tau contributes to endolysosomal dysfunction and neurodegeneration in VCP-related frontotemporal dementia. Brain. 2024 Mar 1;147(3):970–9.

37. Juanes AM, Vogels T, Hertog SK, Loos M, Lutjohann D, Giera M, van der Kant R. Characterization of the brain lipidome associated with frontotemporal lobar degeneration MAPT P301S mutation. J Lipid Res. 2025 Nov 27:100952.

38. Zou M, Li R, Wang JY, et al. Association analyses of variants of SIPA1L2, MIR4697, GCH1, VPS13C, and DDRGK1 with Parkinson’s disease in East Asians. Neurobiol Aging. 2018 Aug;68:159 e7–e14.

39. Andres-Alonso M, Ammar MR, Butnaru I, et al. SIPA1L2 controls trafficking and local signaling of TrkB-containing amphisomes at presynaptic terminals. Nat Commun. 2019 Nov 29;10(1):5448.

40. Sogabe S, Nakano H, Ogasahara Y, et al. Regulation of MCCC1 expression by a Parkinson’s disease-associated intronic variant: implications for pathogenesis. J Hum Genet. 2025 Jul;70(7):371–4.

41. Dominguez-Bautista JA, Klinkenberg M, Brehm N, et al. Loss of lysosome-associated membrane protein 3 (LAMP3) enhances cellular vulnerability against proteasomal inhibition. Eur J Cell Biol. 2015 Mar-Apr;94(3-4):148–61.

42. Steger M, Tonelli F, Ito G, et al. Phosphoproteomics reveals that Parkinson’s disease kinase LRRK2 regulates a subset of Rab GTPases. Elife. 2016 Jan 29;5.

43. Pihlstrom L, Rengmark A, Bjornara KA, et al. Fine mapping and resequencing of the PARK16 locus in Parkinson’s disease. J Hum Genet. 2015 Jul;60(7):357–62.

44. MacLeod DA, Rhinn H, Kuwahara T, et al. RAB7L1 interacts with LRRK2 to modify intraneuronal protein sorting and Parkinson’s disease risk. Neuron. 2013 Feb 6;77(3):425–39.

