## Supplementary Figures for "Genome-wide association studies identify genetic determinants of synucleinopathy biomarkers"

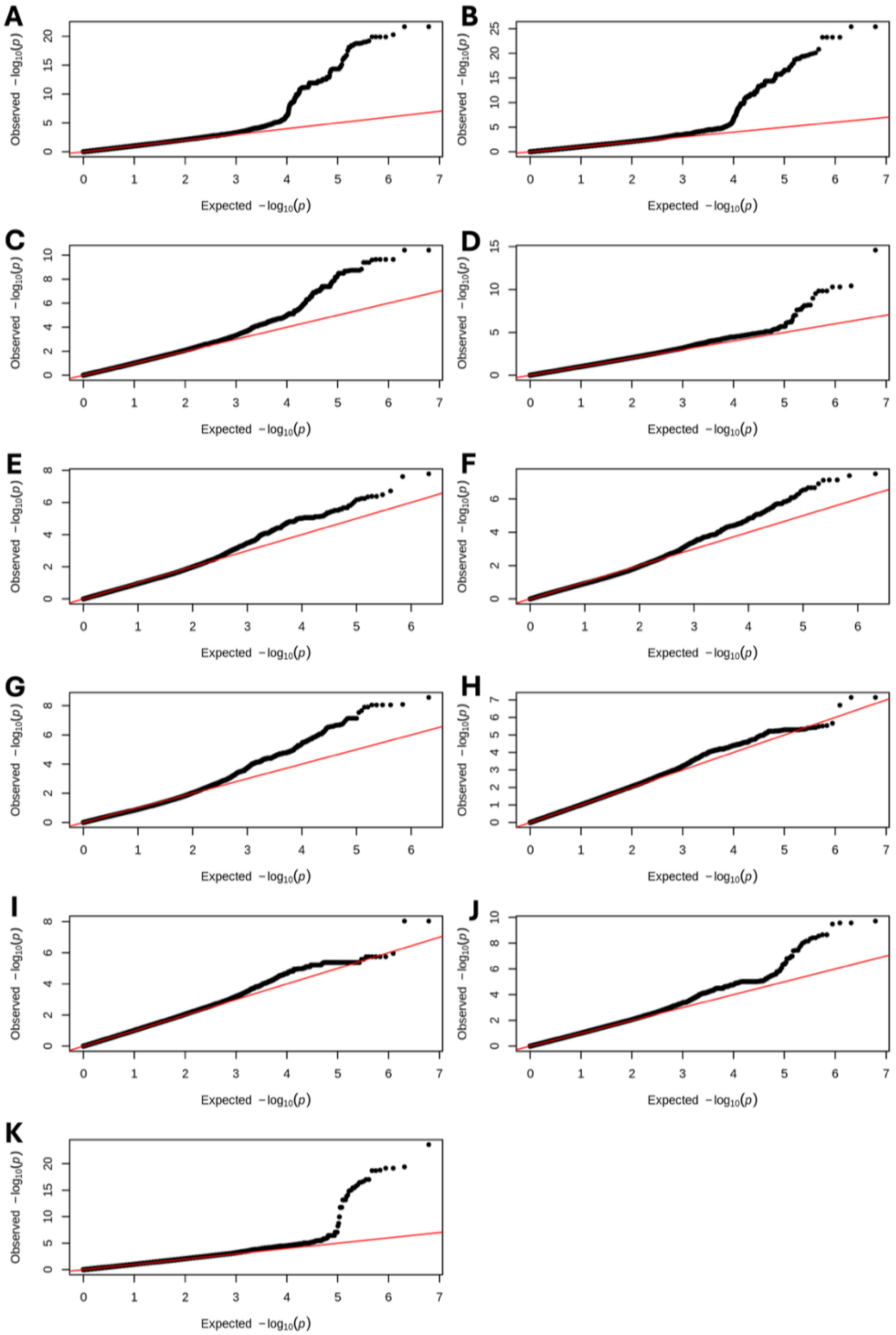


**Supplementary Fig 1.** QQ plots of log-adjusted p-values from A) urine BMP 2,2'-di-22:6, B) urine BMP total di-22:6 BMP, C) urine BMP total di-18:1, D) CSF Ab, E) CSF ceramide C22, F) CSF ceramide C23, G) CSF ceramide C24, H) CSF pTau, I) CSF tTau, J) CSF Ab/pTau, and K) CSF Ab/tTau.

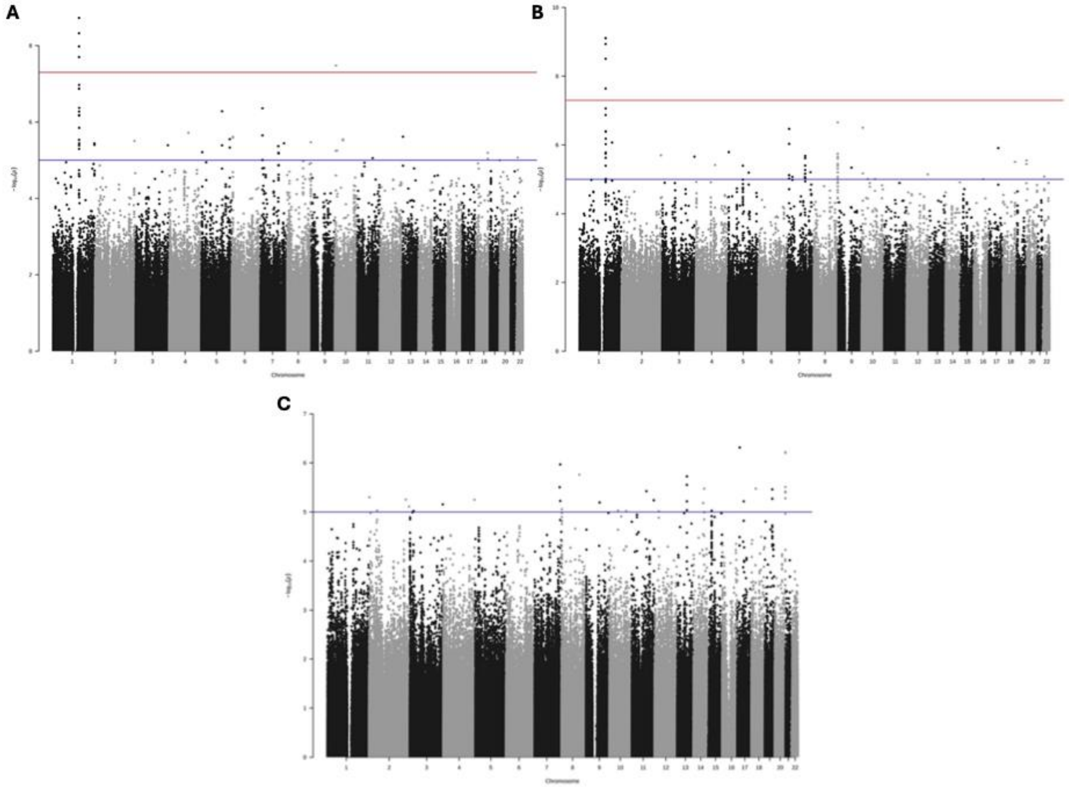


**Supplementary Figure 2.** Manhattan plots of log-adjusted p-values at each genomic position after additional LRRK2 G2019S adjustment for urine BMP A) 2,2'-di-22:6, B) total di-22:6, and C) total di-18:1 isoforms.

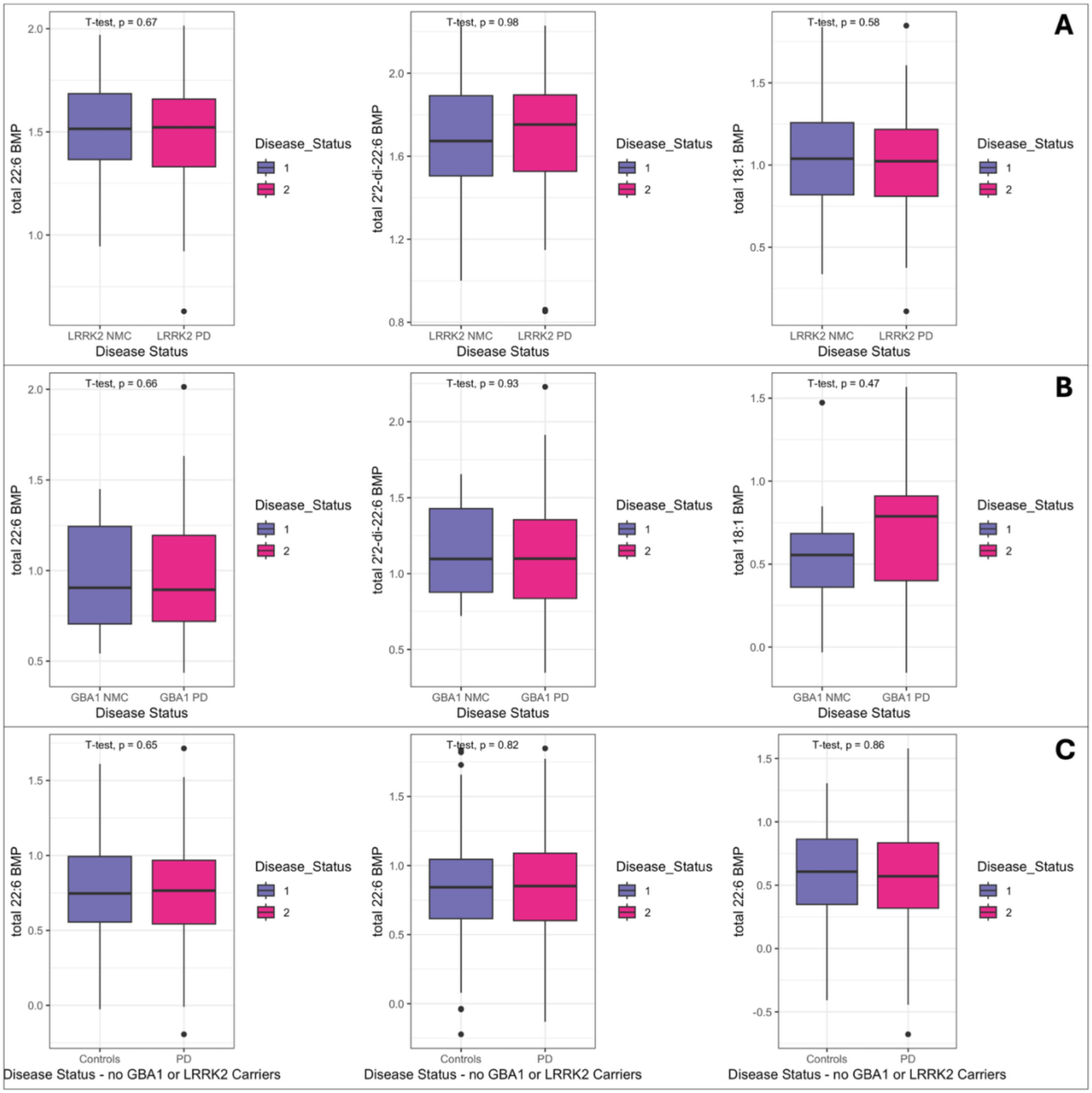


**Supplementary Figure 3.** Histograms for urine BMP isoform levels in the PPMI cohort comparing A) *LRRK2* G2019S carriers with PD to non-manifesting controls, B) *GBA1* N370S carriers with PD to non-manifesting controls, and C) PD patients and controls without *LRRK2* G2019S and *GBA1* N370S mutations.
